## Supplemental Information for "Estimating individual risks of COVID-19-associated hospitalization and death using publicly available data"

**Table 1. Daily contact rates by age and setting.** Contact rates are summed from country and age-specific contact matrixes for the United States supplied in: Prem K, Cook AR, Jit M (2017) Projecting social contact matrices in 152 countries using contact surveys and demographic data. PLoS Comput Biol 13(9): e1005697. https://doi.org/10.1371/journal.pcbi.1005697

| Age | Home | Work | School | Other | Non-home | Total |
|---|---|---|---|---|---|---|
| 20 - 29 Years | 2.77 | 5.39 | 0.94 | 5.55 | 11.88 | 14.66 |
| 30 - 39 Years | 3.20 | 6.09 | 0.80 | 4.22 | 11.10 | 14.30 |
| 40 - 49 Years | 3.18 | 5.92 | 1.11 | 3.47 | 10.49 | 13.67 |
| 50 - 59 Years | 3.33 | 4.61 | 1.70 | 4.22 | 10.53 | 13.86 |

**Table 2. Estimates of the case hospitalization and case fatality ratios.** Computed from US CDC surveillance case report data including case report dates from June 16 to September 15, 2020. Computed from: U.S. Centers for Disease Control and Prevention. COVID-19 Case Surveillance Public Data. Available at: https://data.cdc.gov/Case-Surveillance/COVID-19-Case-Surveillance-Public-Use-Data/vbim-akqf

| Age | Cases | Hospitalizations | Deaths | CHR | CFR |
|---|---|---|---|---|---|
| 20 - 29 Years | 627819 | 10385 | 285 | 1.65% | 0.05% |
| 30 - 39 Years | 478492 | 14998 | 702 | 3.13% | 0.15% |
| 40 - 49 Years | 426258 | 20251 | 1580 | 4.75% | 0.37% |
| 50 - 59 Years | 389910 | 29134 | 3776 | 7.47% | 0.97% |

**Table 3. Cumulative COVID-19 associated hospital admissions rates per 100,000 people during the period June 16 to September 15, 2020**. Computed from US Centers for Disease Control and Prevention COVID-NET program data on laboratory-confirmed Covid-19- Associated Hospitalizations. Available at: https://gis.cdc.gov/grasp/COVIDNet/COVID19_3.html

| Age | Period cumulative hospitalization rate |
|---|---|
| 18-29 years | 44.4 |
| 30-39 years | 59.2 |
| 40-49 years | 78.5 |
| 50-64 years | 109.1 |

**Table 4. Cumulative COVID-19 associated mortality rates per 100,000 people during the period June 16 to September 15, 2020.** Computed from: U.S. Centers for Disease Control and Prevention National Center for Health Statistics Provisional COVID-19 Death Counts by Sex, Age, and Week. Available at: https://data.cdc.gov/NCHS/Provisional-COVID-19-Death-Counts-by-Sex-Age-and-W/vsak-wrfu

| Age | Period deaths | Period cumulative mortality rate |
|---|---|---|
| 15-24 years | 205 | 0.48 |
| 25-34 years | 668 | 1.46 |
| 35-44 years | 1813 | 4.39 |
| 45-54 years | 4499 | 10.81 |
| 55-64 years | 10428 | 24.67 |

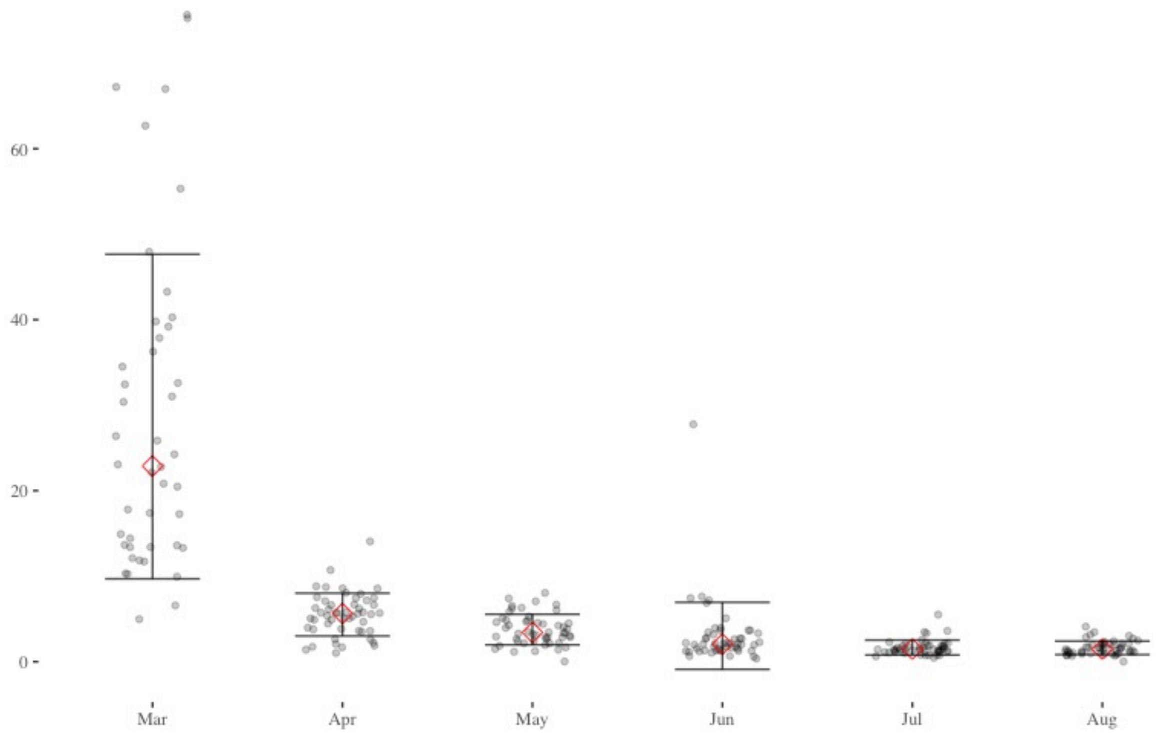


**Fig 1. Estimates of the state-specific case fatality ratio by month.** Computed from: U.S. Centers for Disease Control and Prevention data. United States COVID-19 Cases and Deaths by State over Time. Available at: https://data.cdc.gov/Case-Surveillance/United-States-COVID-19-Cases-and-Deaths-by-State-o/9mfq-cb36.
